# Imaging protocol, feasibility and reproducibility of cardiovascular phenotyping in a large tri-ethnic population-based study of older people: the Southall and Brent Revisited (SABRE) study

**DOI:** 10.1101/2020.08.17.20176305

**Authors:** Lamia Al Saikhan, Muath Alobaida, Anish Bhuva, Nish Chaturvedi, John Heasman, Alun D. Hughes, Siana Jones, Sophie Eastwood, Charlotte Manisty, Katherine March, Arjun K Ghosh, Jamil Mayet, Ayodipupo Oguntade, Therese Tillin, Suzanne Williams, Andrew Wright, Chloe Park

## Abstract

**Background:** People of South Asian and African Caribbean ethnicities living in UK have a high risk of cardiometabolic disease. Limited data exist regarding detailed cardiometabolic phenotyping in this population. Methods enabling this are widely available, but the practical aspects of undertaking such studies in large and diverse samples are seldom reported.

**Methods:** The Southall and Brent Revisited (SABRE) study is the UK’s largest tri-ethnic longitudinal cohort. Over 1400 surviving participants (58-85 y) attended the 2^nd^ study-visit (2008-2011) during which comprehensive cardiovascular phenotyping, including 3D-echocardiography (3D-speckle-tracking (3D-STE)), computed tomography, coronary artery calcium scoring, pulse-wave velocity, central blood pressure, carotid artery ultrasound and retinal imaging were performed. We describe the methods used with the aim of providing a guide to their feasibility and reproducibility in a large tri-ethnic population-based study of older people.

**Results:** Conventional echocardiography and all vascular measurements showed high feasibility (>90% analyzable of clinic-attendees). 3DE and 3D-STE were less feasible in this age group (76% 3DE acquisition feasibility and 38% 3D-STE feasibility of clinic-attendees). Intra- and inter-observer variabilities were excellent for most of conventional and advanced echocardiographic measures. The test-retest reproducibility was good-excellent and fair-good for conventional and advanced echocardiographic measures, respectively, but lower than when re-reading the same images. All vascular measures demonstrated excellent or fair-good reproducibility.

**Conclusions:** Detailed cardiovascular phenotyping is feasible and reproducible in an ethnically diverse population. The data collected will lead to a better understanding of why people of South Asian and African Caribbean ancestry are at elevated risk of cardiometabolic diseases.

## 1 Introduction

People of South Asian and African Caribbean descent are known to experience an increased burden of diabetes and cardiovascular disease (CVD) compared with people of European ancestry, although why remains unclear (1). The Southall and Brent Revisited study (SABRE) was designed to address this question by establishing the relationships between risk factors, subclinical disease and adverse clinical outcomes and mortality in a longitudinal cohort (2).

Methods enabling detailed cardiometabolic phenotyping are widely available but the practicalities undertaking phenotyping in large and diverse study populations are seldom reported (3). We describe the clinical cardiovascular imaging acquisition and analysis methods performed on over 1400 participants in SABRE aged between 58 and 85 years in the second-wave of follow-up with the aim of providing information on the feasibility and reproducibility of detailed measures of cardiovascular structure and function in a community-based study of older people.

## 2 Methods

### 2.1 SABRE Cohort

The design of SABRE has been described in detail previously (2). Briefly, all traceable surviving participants from the baseline studies conducted in 1988 to 1991 were invited to attend the second-wave of follow-up at St Mary’s Hospital, London. Of 3410 survivors, 1438 (42%) participants attended the clinic between 2008 and 2011 (69.6±6 y). Participants attended for clinical investigations as listed in **Table-S1**. They also completed a health and lifestyle questionnaire and were asked for consent to primary care medical record review. All participants gave written informed consent and the study was approved by the Local Research Ethics Committee.

### 2.2 Echocardiographic Measurements

The SABRE study employed standard and advanced echo techniques characterized (1) left ventricular (LV) structure and geometry, (2) LV systolic function, (3) LV dyssynchrony, (4) LV diastolic function, and (5) left atrial(LA) structure. The incremental value of LV mechanics by 3D-speckle-tracking echocardiography (3D-STE) in predicting adverse outcomes is of particular interest, considering that SABRE is the first multi-ethnic longitudinal study to capture 3D-STE measures.

#### 2.2.1 Data Acquisition Protocol

All participants underwent a transthoracic-echocardiographic examination using a Phillips iE33 equipped with a S5-1 phased array ultrasound transducer for 2D and Doppler imaging and a matrix array (X3-1) transducer for 3D-data acquisition. A brief outline of the SABRE echocardiography-imaging protocol is listed in **Table-1**. The examination was performed by two experienced sonographers in accordance with ASE guidelines (4). Echocardiographic measures were obtained from the parasternal long-axis (PLAX), parasternal short-axis and apical four-, two- and five-chamber views. Optimal images were attained prior to recording and harmonic imaging was used when fundamental B-mode imaging was unsatisfactory. An adequate quality of ECG signal was ensured throughout the examination. 3-5 and 10 cardiac-cycles were recorded for 2D-images and spectral-Doppler imaging, respectively.

**Table.1.** SABRE echocardiographic imaging protocol.

| ***Parasternal position*** |  |
| --- | --- |
| Parasternal long axis | - 2D imaging of LV<br>▪ 3 cardiac-cycles<br>- 2D imaging, zoomed on LVOT<br>- M-mode imaging focused on LA<br>▪ 10 cardiac-cycles with a sweep speed of 50 msec<br>- Color Doppler of mitral and aortic valves |
| Parasternal short axis | - 2D imaging<br>▪ 5 cardiac-cycles |
| - Mitral valve level |  |
| - Papillary muscle level |  |
| - LV apex |  |
| ***Apical position*** |  |
| Apical four-chamber view | - 2D imaging<br>- 2D imaging, focused/zoomed on LV<br>▪ 5 cardiac-cycles<br>- PW Doppler of mitral inflow<br>▪ 2-5 mm SV<br>▪ 50-100 mm/sec sweep speed<br>▪ 10 cardiac-cycles<br>- Tissue Doppler imaging of septal and lateral mitral valve annulus<br>- 3D full-volume of acquisition of LV<br>▪ Four sub-volumes over 4 cardiac-cycles |
| Apical five-chamber view | - 2D imaging<br>- Color Doppler of aortic valve<br>- CW Doppler of trans-aortic flow<br>▪ 10 cardiac-cycles |
| Apical two-chamber view | - 2D imaging focused/zoomed on LV<br>▪ 5 cardiac-cycles<br>- Tissue Doppler imaging of inferior wall |

CW, continuous wave; LA, left atrium; LV, left ventricle; LVOT, LV outflow tract; PW, pulse wave.

2D-image optimization included depth, sector width and gain setting. Harmonic imaging was used when needed to ensure clear visualization of endocardial borders for quantitative analysis including volumes and 2D-STE analysis. Parallel ultrasound beam orientation to the blood flow of interest was ensured for all Doppler acquisitions with a sample volume size 2-5 mm for PW Doppler and a sweep speed between 50-100 mm/sec.

For 3D-imaging, a LV full-volume dataset was obtained from the apical position. Depth and sector width setting and gain were adjusted as needed (5). 4 sub-volumes acquired over 4 cardiac-cycles during held respiration were obtained in the wide-angled (93º×80º) acquisition mode. The presence of an acceptable ECG signal was checked before accepting the loop. Scans were stored on a secure fileserver in DICOM or native format.

#### 2.2.2 Data Analysis

All conventional echocardiographic analyses were performed on the ultrasound machine during the clinic visit using Philips QLAB software 7.0, averaging 3 measurements. 2D-STE analysis was performed offline using Philips QLAB software 8.1 Tissue Motion Quantification Advanced (TMQA). 3DE analysis was performed using Philips QLAB software 7.0 with 3DQ (Cardiac 3D-Quantification) for LV mass and 3DQ Advanced for LV volumes. 3D-STE LV myocardial deformation analysis was performed using TomTec 4D LV-Analysis (TomTec Imaging Systems, Munich, Germany).

Measured echocardiographic parameters are listed in **Table-2**. LV and LA dimensions, outflow-tract diameter and LV wall thickness from 2D-guided M-mode were measured from the PLAX from which LV mass was calculated, following the ASE recommendations (6). LV hypertrophy (LVH) based on conventional-echocardiography-derived LV mass was defined as LV mass indexed to body surface area (BSA)(7) > 115g/m^2^ in men and 95g/m^2^ in women (6). Relative wall thickness (RWT) was calculated (6). LV geometry was categorized as follows: normal if RWT≤0.42 and no LVH; LV remodeling if RWT>0.42 and no LVH; LV concentric hypertrophy if RWT>0.42 and LVH; and LV eccentric hypertrophy if RWT≤0.42 and LVH (6). LV volumes from conventional echo were calculated by the Teichholz formula using the linear dimensions from which LV ejection fraction (LVEF) was derived (6). Midwall fractional shortening was also calculated (8, 9). Tissue-Doppler analysis of lateral and septal mitral annulus motion was performed and peak longitudinal systolic velocity (s′), and peak early and late mitral annular relaxation velocities (e′, a′) were calculated. Mitral inflow early(E-wave) and late (A-wave) diastolic velocities were measured by PW Doppler with a sample volume placed at the tip of mitral valve leaflets (10). E/e′ was calculated as an index of LV filling pressure (10).

**Table.2.**
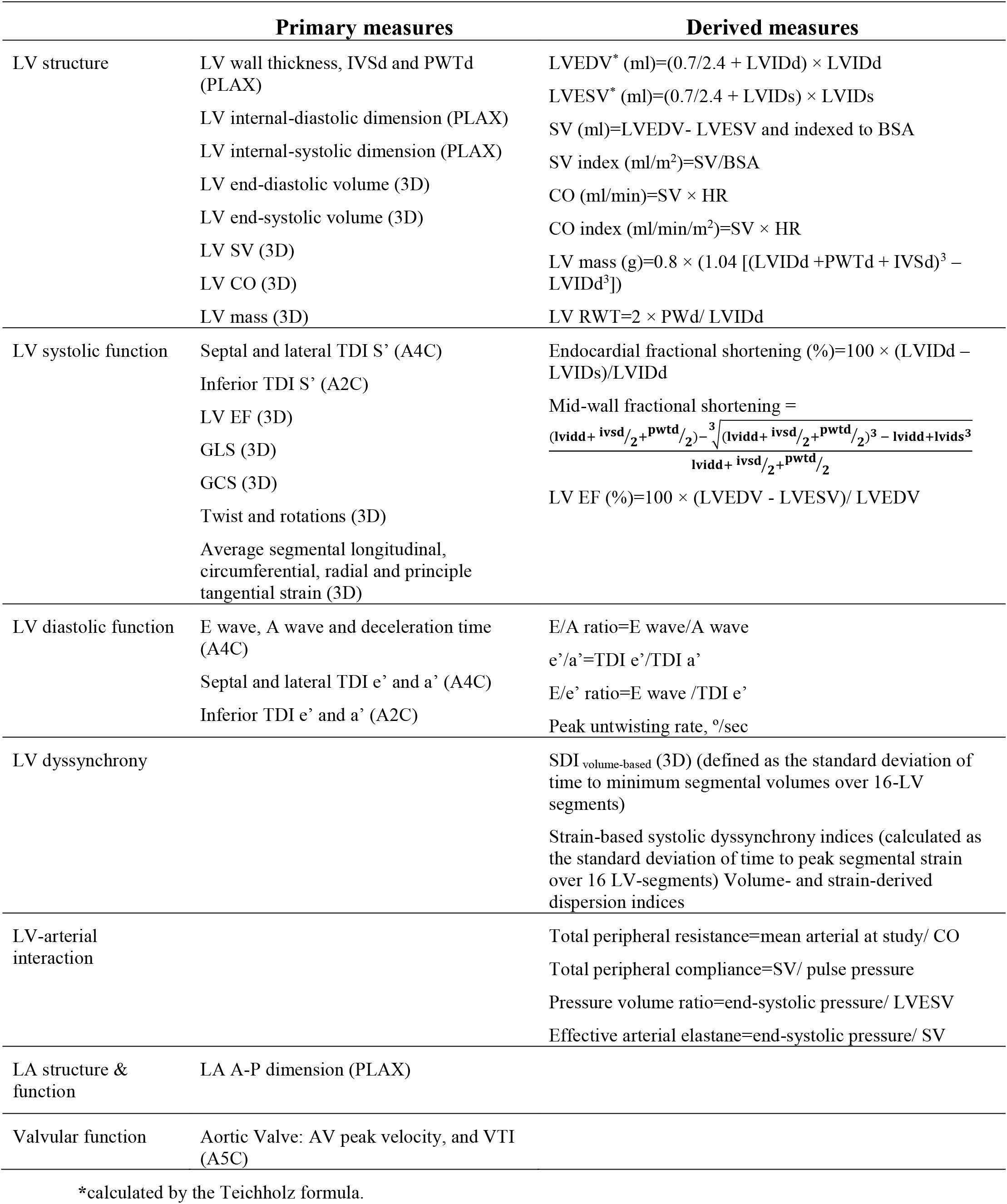
SABRE echocardiographic measures.

3DE LV mass and volumetric analyses were performed offline according to a pre-developed protocol. Only 3D-datasets of the highest quality free of stitching artefact were used. All views were optimized to ensure that the LV was not foreshortened. For LV mass, the endocardial and epicardial boundaries of the apical four- and two-chamber views were traced at end-diastole (ED), the first frame after mitral valve closure, and end-systole (ES), the first frame after aortic valve closure. Cardiac 3DQ calculated LV mass using the biplane method of discs. LVH based on 3DE-derived LV mass was determined using the 95^th^ upper percentiles of indexed LV mass reported by the Multi-Ethnic Study of Atherosclerosis study (11). Cardiac 3DQ Advanced was used to measure 3D LV volumes without the geometrical assumptions of 2D methods. Five reference points were selected on the LV in two orthogonal views in each ED and ES frames. The software then automatically tracked the endocardium from frame to frame throughout the cardiac-cycle to calculate the volumes and LVEF and constructed a 3D shell of the LV and global and regional volumetric waveforms. Manual adjustments were performed when needed.

LV advanced myocardial mechanics were assessed according to a pre-specified protocol. Acceptable image quality was defined as follows:

1. **Good** = clearly visualized endocardium in all 16 segments in ED and ES frames.
2. **Fair** = not clearly visualized endocardium in ≤2 segments or when there were just minor artefacts present e.g. apical noise
3. **Adequate** = not clearly visualized in endocardium in ≤6 segments
4. **Poor** = clearly visualized endocardium in > 6 segments in ED or ES frames, but the endocardium was still be tracked with confidence throughout the cardiac-cycle using the adjacent segments as a reference

Unacceptable image quality was defined as follows:

1. Major stitching artefacts preventing reliable tracking of the endocardium
2. Unacceptable visualization of the endocardial boundaries
3. Multiple segments (≥4 = ≥25%) of the LV wall being outside of the image sector

The identity of non-visible segments were recorded in a spreadsheet during analysis as some regions are known to be less feasible, such as the apex, anterior and anterolateral segments (12). A 16-segment model was used and LV segments were scored as 0 if non-visible and 1 if visible (6).

LV endocardial borders at ED were automatically tracked and manual adjustments were performed when needed (12, 13). LV volumes including LVEF and systolic dyssynchrony index (SDI), global longitudinal (GLS) and circumferential (GCS) strains, and average peak segmental longitudinal, circumferential, radial and principle tangential strains, and twist and rotational indices were calculated by the software. Additional measures were derived: 1)strain-based systolic dyssynchrony indices, calculated as the standard deviation of time to peak segmental strain over 16 LV-segments normalized to cardiac-cycle length, and 2)volume- and strain-derived dispersion (Di) indices, defined as difference between minimum and maximum time to peak of a measure over 16-LV segments normalized to cardiac-cycle length (14).

### 2.3 Vascular Measurements

#### 2.3.1 Pulse Wave Velocity

Pulse Wave Velocity (PWV) is a gold standard measure of arterial stiffness (15). The assessment of PWV relies on the determination of pulse transit time (TT), defined as the time the pulse wave takes to travel from one point to another over an arterial segment of length (L).

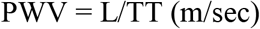

The distance from the sternal notch to the common femoral artery (mm) at the inguinal ligament was measured using a measuring tape while the participants lay in a supine position.

A PT2000 (Micro Medical Ltd, Kent) was used to acquire 10 sequential blood velocity waveforms at the carotid and femoral artery using a 4MhZ CW directional Doppler probe with a zero-crossing detector and the ECG R-wave as a time reference. At least two windows of good quality flow traces were recorded.

#### 2.3.2 Central Blood Pressure and Pulse Wave Analysis

Aortic (central) blood pressure (BP) differs from brachial (peripheral) BP and may be a superior predictor of cardiovascular events (16). Radial artery applanation tonometry was assessed using the SphygmoCor device (Atcor, Sydney, Australia) to record at least six cardiac-cycles of central BP waveforms, estimated after calibrating the device to resting brachial systolic and diastolic BP (17).

#### 2.3.3 Coronary Artery Calcification

Computed tomography (CT) coronary artery calcium scores (CACS) were measured in those without stents using a Philips MX 8000 IDT 64 Detector scanner (2.5 mm slice thickness and 25 mm increments). Images were transferred to a workstation for offline analysis (HeartBeat CS) by an experienced radiographer blinded to participant identity. Areas of calcification were assigned to the 4 main territories of the coronary arteries: left main, left anterior descending, circumflex and right coronary artery.

#### 2.3.4 Common Carotid Intimal Medial Thickness

Vascular ultrasound of the left carotid artery was performed using Phillips iE33 equipped with a linear-array transducer (L11-3). Defult settings (FR = 50, Depth = 2.5 cm, Gain = 62 dB, C = 51dB, P = Low and Pen) were used, but adjustments were allowed to optimise image quality. A cine loop of at least 5 cycles at 3 angles (lateral, posterior and anterior) as well as one still image captured at the R-wave for each angle were acquired. Data were analysed offline using the AMS-II (v1.1364) (18). The region of interest was defined as 1 cm proximal to the carotid bulb with the analysed image chosen to ensure optimal quality of the far wall. Modifications of the detected borders or length were performed when necessary. Carotid plaque was defined as the presence of focal wall thickening at least 50% greater than that of the surrounding vessel wall or as a focal region with cIMT greater than 1.5 mm protruding into the lumen distinct from the adjacent boundary (19). Plaque characteristics including area, gray-scale median and percentage white were measured using the AMS-II software.

#### 2.3.5 Retinal Imaging

Fundus imaging of both eyes was performed in people without glucoma or any condition that prevented adequate retinal imaging using a Zeiss FF450+ fundus-camera and an Oscar 510CCCD after the administration of eye drops (1% tropicamide and 2.5% phenylephrine). Refraction was measured using a Nidek AR-310 auto-refractometer. Retinopathy was defined according to the NHS Diabetic Eye Screening Programme, and quantitative measures of the retinal microvasculature were performed as described (20). Feasibility and reproducibility have been reported previously (20).

### 2.4 Quality Control and Reproducibility Analyses

All clinic staff underwent training and accreditation and data quality was monitored regularly throughout the study. To quantify repeatability and reproducibility of echo measurements, two sonographers independently analyzed the same echocardiograms in subsets of randomly selected participants (*n* = 10 for conventional echocardiographic measures, and *n* = 20 for 3DE measures) with an interval of at least two weeks to avoid recall bias and blinded to the first observer measurements. Test-retest reproducibility of conventional and 3D-STE analysis was performed in selected participants who agreed to re-attend the clinic four to eight weeks after the first visit. The repeatability and reproducibility of vascular measurements was performed following the same approach in at least 10 randomly selected subjects.

#### 2.4.1 Statistical Analysis

Repeated measurements were assessed by intraclass correlation coefficient (ICC) calculated by linear mixed modelling. Reliability was classified as follows: ICC< 0.4 = poor, 0.4≥ICC< 0.75 = fair to good, and ICC≥0.75 = excellent (21). Agreement was measured by Bland Altman (BA) analysis and reported as the mean difference with 95% limits of agreement. All BA graphs of the intra-observer (repeatability) and inter-observer (reproducibility) reliability of the echocardiographic and vascular measures are shown in the online supplementary material. All analyses were performed in Stata 15.1(StataCorp LLC, USA).

## 3 Results

### 3.1 Feasibility

Brief characteristics of the population are summarized in **Table-3**.

**Table.3.** Characteristics of SABRE participants (Visit-2)

| ***n*=1438** | **Mean±SD** |
| --- | --- |
| Age, y | 69.7±6.2 |
| Male, n(%) | 1092(75.9) |
| Europeans/South Asians/African Caribbeans, n(%) | 684(47.6)/522(36.3)/232(16.1) |
| Height, cm | 168.0±8.6 |
| Weight, kg | 78.0±15.0 |
| Body mass index, kg/m<sup>2</sup> | 27.6±4.8 |
| Hypertension, n(%) | 965(67.1) |
| Diabetes, n(%) | 451(31.4) |
| Coronary heart disease, n(%) | 362(25.2) |
| Stroke, n(%) | 81(5.6) |
| Disability, n(%) | 569(39.8) |
| Smoking status (never/ex/current), n(%) | 810(56.7)/529(37.0)/90(6.3) |

Disability was defined as a get up and go test time ≥12 sec.

#### 3.1.1 Echocardiography

2D, spectral- and tissue-Doppler echocardiography was analyzable in up to 95% of clinic attendees (**Table-4, Figure-1A**). 3DE was not acquired in participants with atrial fibrillation or poor image quality, and there was a small subset of participants who had attended the clinic before the 3D probe became available; these were excluded from the denominator when feasibility was estimated. 3DE was acquired in 76% of all clinic attendees and, using QLAB, 924 (66%) had successful volumetric analysis and 905 (65%) had LV mass calculated. The difference in these numbers reflects difficulties in tracking the epicardium compared to the endocardium. 50% of those who had 3DE-datasets had 3D deformation measurements by Tomtec. Reasons for non-analysis included unacceptable images for strain analysis, stitching artefacts and low frame rate. The feasibility of 3D-STE was 38% of all clinic attendees.

**Figure 1.**
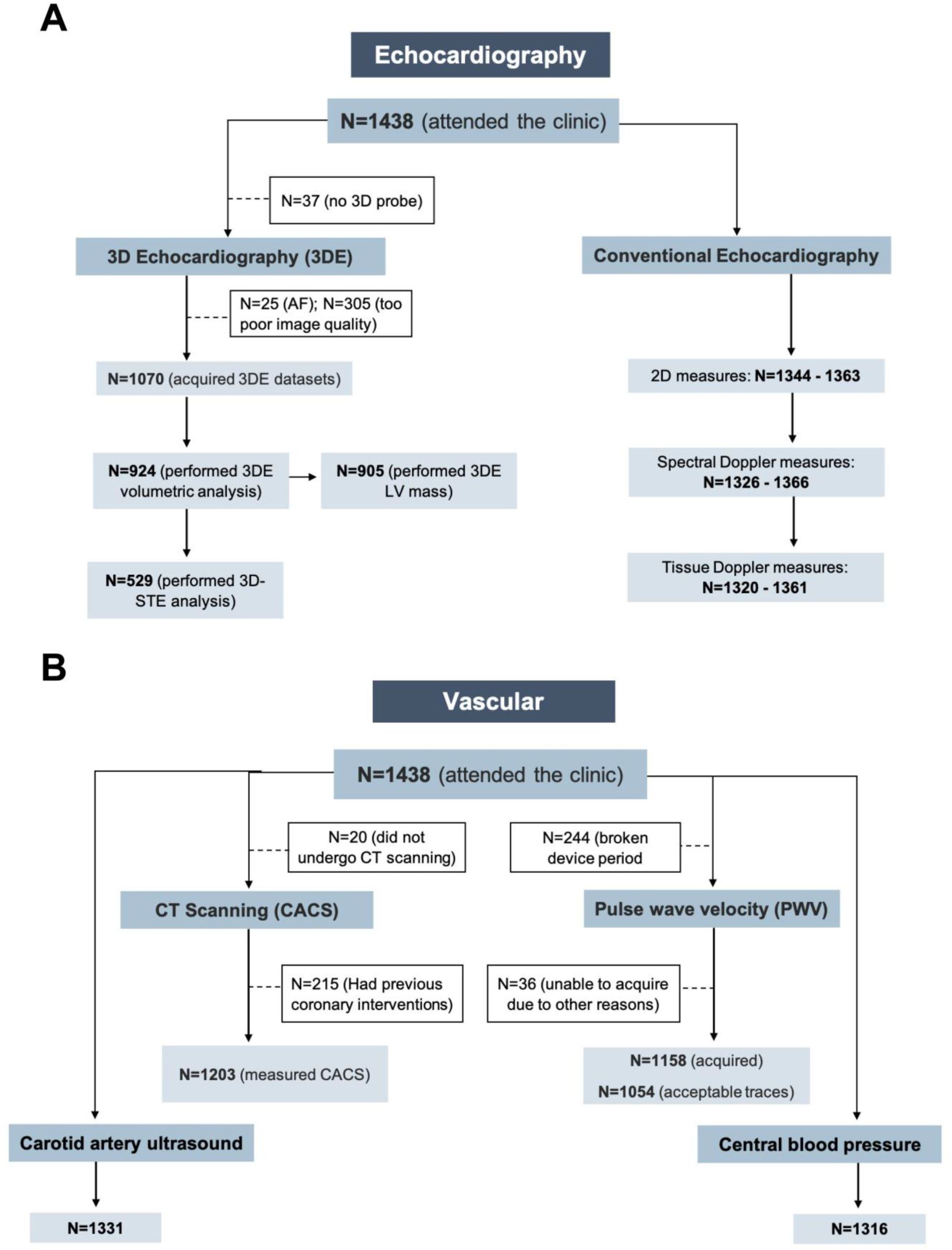
Feasibility of the cardiovascular measurements in SABRE study. AF, atrial fibrillation; CACS, coronary artery calcium score; CT, computed tomography.

**Table.4.** Feasibility of the cardiovascular measures in 1438 SABRE participants.

| **2D** |  |
| --- | --- |
| LVIDd | 1354(94%) |
| LVIDs | 1352(94%) |
| IVSd | 1354(94%) |
| IVSs | 1352(94%) |
| PWd | 1354(94%) |
| PWs | 1353(94%) |
| LA diameter | 1344(93%) |
| LVOT diameter | 1363(95%) |
| **Spectral-Doppler** |  |
| AV VTI | 1355(94%) |
| AV max velocity | 1358(94%) |
| E wave | 1366(95%) |
| A wave | 1326(92%) |
| Deceleration time | 1360(95%) |
| **Tissue-Doppler** |  |
| e' septal | 1359(95%) |
| a' septal | 1320(92%) |
| s' septal | 1362(95%) |
| e' lateral | 1360(95%) |
| a' lateral | 1321(92%) |
| s' lateral | 1361(95%) |
| E/e' | 1337(93%) |
| **3DE** |  |
| QLAB EF, EDV, ESV | 924(66%) |
| QLAB LV mass | 905(65%) |
| **3D-STE\*** |  |
| GLS, GCS | 529(50%) |
| Twist and rotations | 529(50%) |
| **Vascular\*** |  |
| cIMT | 1331(92.5%) |
| Central SBP and DBP | 1316(91.5%) |
| AIx, | 1316(91.5%) |
| Total CACS | 1203(83.7%) |
| PWV | 1054(91%) |

\*see text in the manuscript for details.

#### 3.1.2 Vascular Measurements

Central BP, cIMT and CACS showed high feasibility with good quality measurements achievable in 92%, 93% 84% of clinic attendees respectively (**Figure-1B; Table-4**). PWV was not collected in 244 participants due to device failure, excluding this reason for missing data gave a feasibility of 97%. CACS was measurable in all participants who underwent CT scanning, 15% were ineligible due to stent placement, and 1% of eligible individuals did not undergo CT scanning either due to refusal or unavailability of the scanner.

### 3.2 Repeatability and Reproducibility

#### 3.2.1 Echocardiography

Intra-observer (repeatability) and inter-observer (reproducibility) reliability of conventional echocardiographic measures was good or excellent for both read-reread and test-retest studies (**Table-5**).

**Table.5.** Reproducibility of conventional echocardiographic measures.

| Bland & Altman Analysis |  |  |  |  |  |  |
| --- | --- | --- | --- | --- | --- | --- |
| Mean Diff (95% LOA), ICC |  |  |  |  |  |  |
|  | Intra-observer(n=10) |  | Inter-observer(n=10) |  | Test-retest(n=36) |  |
| 2D measures |  |  |  |  |  |  |
| LA dimension, cm | -0.02(-0.17, 0.13) | 0.97 | -0.06(-0.25, 0.12) | 0.98 | -0.001(-0.23, 0.23) | 0.98 |
| IVSd, cm | -0.01(-0.11, 0.08) | 0.91 | -0.07(-0.33, 0.17) | 0.85 | -0.01(-0.16, 0.14) | 0.94 |
| LVIDd, cm | 0.001(-0.23, 0.23) | 0.96 | -0.007(-0.20, 0.18) | 0.97 | 0.001(-0.34, 0.34) | 0.95 |
| PWTd, cm | -0.01(-0.13, 0.10) | 0.96 | -0.03(-0.16, 0.09) | 0.82 | -0.02(-0.25, 0.22) | 0.70 |
| IVSs, cm | -0.007(-0.09, 0.08) | 0.92 | 0.005(-0.09, 0.10) | 0.96 | 0.003(-0.22, 0.23) | 0.92 |
| LVIDs, cm | 0.04(-0.17, 0.26) | 0.97 | 0.02(-0.16, 0.21) | 0.96 | 0.01(-0.43, 0.46) | 0.93 |
| PWTs, cm | -0.04(-0.21, 0.12) | 0.93 | 0.01(-0.30, 0.33) | 0.77 | -0.001(-0.21, 0.21) | 0.88 |
| LVOT diameter, cm | -0.02(-0.14, 0.10) | 0.96 | -0.007(-0.10, 0.09) | 0.96 | 0.01(-0.2, 0.21) | 0.86 |
| Spectral-Doppler measures |  |  |  |  |  |  |
| AV peak velocity, m/s | -3.8(-9.9, 2.3) | 0.97 | 0.001(-0.08, 0.08) | 0.99 | 0.02(-0.25, 0.28) | 0.88 |
| AV VTI, cm | 0.14(-2.8, 3.1) | 0.96 | 0.32(-6.1, 6.8) | 0.87 | 0.11(-7.0, 7.3) | 0.86 |
| E wave, cm/s | 1.5(-4.1, 7.2) | 0.98 | -2.3(-6.7, 2.1) | 0.95 | 0.35(-17.6, 18.3) | 0.84 |
| A wave, cm/s | -0.53(-11.2, 10.2) | 0.91 | -1.4(-4.7, 1.7) | 0.99 | 0.23(-14.7, 15.2) | 0.90 |
| Deceleration time, ms | 10.9(-34.2, 56.0) | 0.82 | 7.4(-18.2, 33.0) | 0.97 | -2.9(-87.1, 81.3) | 0.58 |
| Tissue-Doppler measures |  |  |  |  |  |  |
| s', cm/s | -0.30(-1.4, 0.81) | 0.95 | 0.24(-0.62, 1.1) | 0.95 | -0.20(-1.65, 1.25) | 0.89 |
| e', cm/s | 0.04(-0.9, 1.0) | 0.91 | -0.30(-1.2, 0.58) | 0.98 | -0.43(-3.5, 2.6) | 0.66 |
| a', cm/s | 0.16(-0.79, 1.1) | 0.98 | -0.24(-0.81, 0.33) | 0.99 | -0.24(-3.1, 2.7) | 0.78 |

Data are mean difference (95% limit of agreement (LOA)).
Abbreviations as in table-4 in addition to: ICC, interclass correlation coefficient.

3D volumetric analysis using QLAB software showed excellent reliability for all measures. Mean difference (95% limit of agreement (LOA)) and ICC of the inter-observer difference were −0.06 (−5.5, 5.4) and 0.82 ICC for LVEF (%), −0.1 (−8.2, 8.0) ml and 0.96 ICC for end-diastolic volume, 0.0 (−4.6, 4.6) ml and 0.96 ICC for end-systolic volume, and 1.3 (−7.5, 10.2) g and 0.98 ICC for LV mass. The intra-observer difference for LV mass was −3.5 (−18.2, 11.1) g and 0.96 ICC.

LV volumes, LVEF and LV mass derived from 3D-STE analysis using Tomtec showed excellent repeatability and reproducibility (**Table-6**). Global and averaged peak segmental LV strain measures also showed excellent repeatability and reproducibility, except for longitudinal strain measures, which showed fair to good inter-observer variability. Rotational measures showed fair to good repeatability and reproducibility. All SDIs showed good to excellent repeatability and reproducibility. Dispersion indices also were reproducible apart from circumferential strain-Di, which showed poor repeatability and reproducibility. Overall, reproducibility was better for SDIs than for dispersion indices (**Table-6**).

**Table.6.**
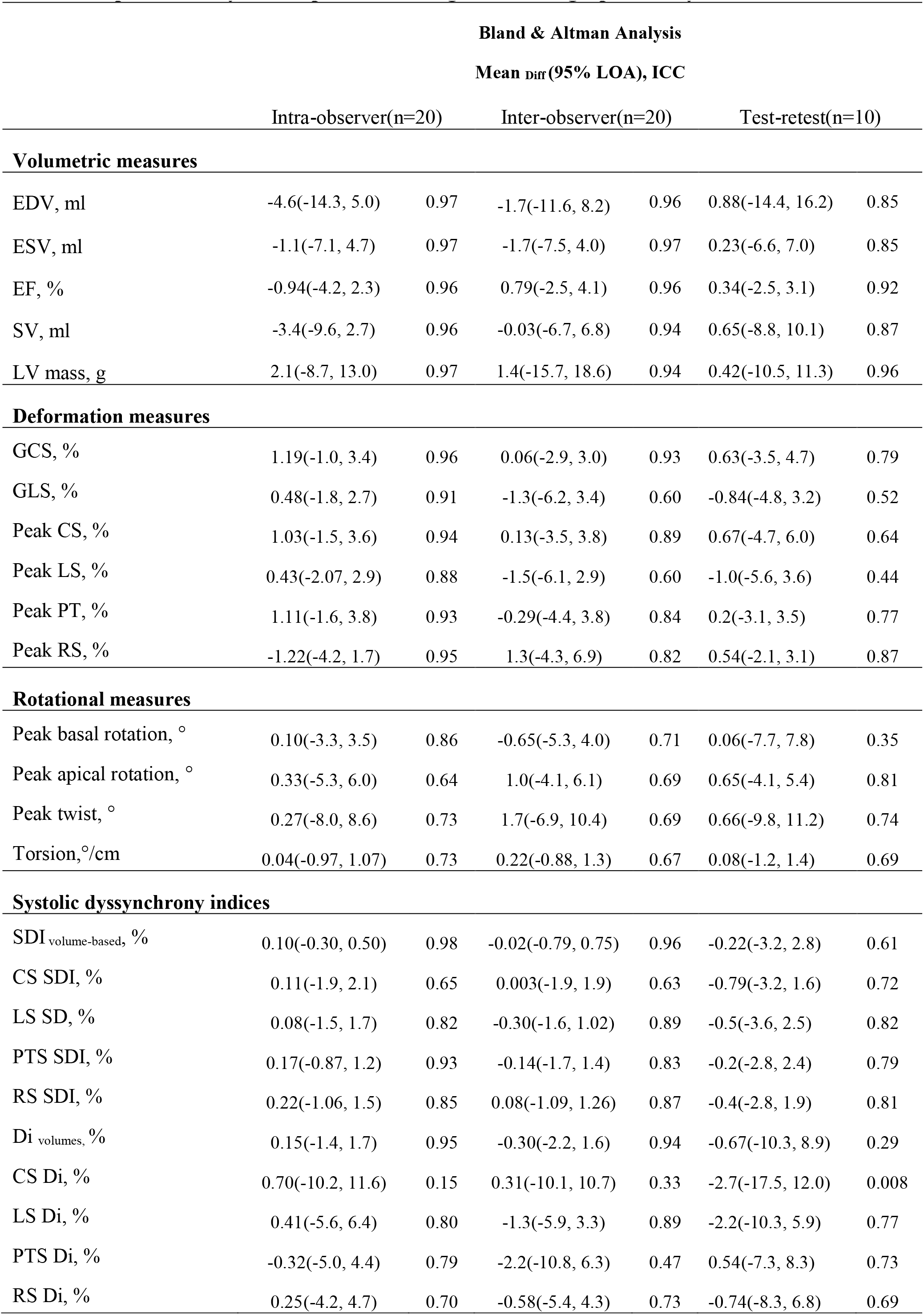
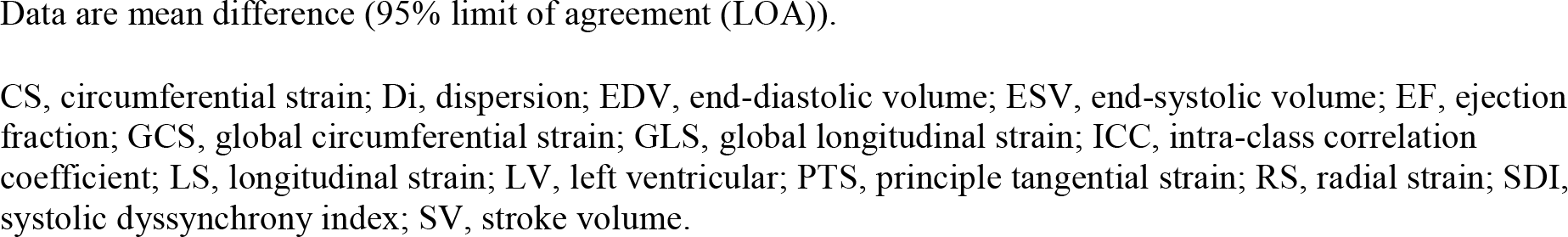
Reproducibility of 3D-speckle tracking echocardiographic analysis.

| **Bland & Altman Analysis** |  |  |  |  |  |  |
| --- | --- | --- | --- | --- | --- | --- |
| **Mean Diff (95% LOA), ICC** |  |  |  |  |  |  |
|  | Intra-observer(n=20) |  | Inter-observer(n=20) |  | Test-retest(n=10) |  |
| **Volumetric measures** |  |  |  |  |  |  |
| EDV, ml | -4.6(-14.3, 5.0) | 0.97 | -1.7(-11.6, 8.2) | 0.96 | 0.88(-14.4, 16.2) | 0.85 |
| ESV, ml | -1.1(-7.1, 4.7) | 0.97 | -1.7(-7.5, 4.0) | 0.97 | 0.23(-6.6, 7.0) | 0.85 |
| EF, % | -0.94(-4.2, 2.3) | 0.96 | 0.79(-2.5, 4.1) | 0.96 | 0.34(-2.5, 3.1) | 0.92 |
| SV, ml | -3.4(-9.6, 2.7) | 0.96 | -0.03(-6.7, 6.8) | 0.94 | 0.65(-8.8, 10.1) | 0.87 |
| LV mass, g | 2.1(-8.7, 13.0) | 0.97 | 1.4(-15.7, 18.6) | 0.94 | 0.42(-10.5, 11.3) | 0.96 |
| **Deformation measures** |  |  |  |  |  |  |
| GCS, % | 1.19(-1.0, 3.4) | 0.96 | 0.06(-2.9, 3.0) | 0.93 | 0.63(-3.5, 4.7) | 0.79 |
| GLS, % | 0.48(-1.8, 2.7) | 0.91 | -1.3(-6.2, 3.4) | 0.60 | -0.84(-4.8, 3.2) | 0.52 |
| Peak CS, % | 1.03(-1.5, 3.6) | 0.94 | 0.13(-3.5, 3.8) | 0.89 | 0.67(-4.7, 6.0) | 0.64 |
| Peak LS, % | 0.43(-2.07, 2.9) | 0.88 | -1.5(-6.1, 2.9) | 0.60 | -1.0(-5.6, 3.6) | 0.44 |
| Peak PT, % | 1.11(-1.6, 3.8) | 0.93 | -0.29(-4.4, 3.8) | 0.84 | 0.2(-3.1, 3.5) | 0.77 |
| Peak RS, % | -1.22(-4.2, 1.7) | 0.95 | 1.3(-4.3, 6.9) | 0.82 | 0.54(-2.1, 3.1) | 0.87 |
| **Rotational measures** |  |  |  |  |  |  |
| Peak basal rotation, ° | 0.10(-3.3, 3.5) | 0.86 | -0.65(-5.3, 4.0) | 0.71 | 0.06(-7.7, 7.8) | 0.35 |
| Peak apical rotation, ° | 0.33(-5.3, 6.0) | 0.64 | 1.0(-4.1, 6.1) | 0.69 | 0.65(-4.1, 5.4) | 0.81 |
| Peak twist, ° | 0.27(-8.0, 8.6) | 0.73 | 1.7(-6.9, 10.4) | 0.69 | 0.66(-9.8, 11.2) | 0.74 |
| Torsion, °/cm | 0.04(-0.97, 1.07) | 0.73 | 0.22(-0.88, 1.3) | 0.67 | 0.08(-1.2, 1.4) | 0.69 |
| **Systolic dyssynchrony indices** |  |  |  |  |  |  |
| SDI<sub>volume-based</sub>, % | 0.10(-0.30, 0.50) | 0.98 | -0.02(-0.79, 0.75) | 0.96 | -0.22(-3.2, 2.8) | 0.61 |
| CS SDI, % | 0.11(-1.9, 2.1) | 0.65 | 0.003(-1.9, 1.9) | 0.63 | -0.79(-3.2, 1.6) | 0.72 |
| LS SD, % | 0.08(-1.5, 1.7) | 0.82 | -0.30(-1.6, 1.02) | 0.89 | -0.5(-3.6, 2.5) | 0.82 |
| PTS SDI, % | 0.17(-0.87, 1.2) | 0.93 | -0.14(-1.7, 1.4) | 0.83 | -0.2(-2.8, 2.4) | 0.79 |
| RS SDI, % | 0.22(-1.06, 1.5) | 0.85 | 0.08(-1.09, 1.26) | 0.87 | -0.4(-2.8, 1.9) | 0.81 |
| Di<sub>volumes</sub>, % | 0.15(-1.4, 1.7) | 0.95 | -0.30(-2.2, 1.6) | 0.94 | -0.67(-10.3, 8.9) | 0.29 |
| CS Di, % | 0.70(-10.2, 11.6) | 0.15 | 0.31(-10.1, 10.7) | 0.33 | -2.7(-17.5, 12.0) | 0.008 |
| LS Di, % | 0.41(-5.6, 6.4) | 0.80 | -1.3(-5.9, 3.3) | 0.89 | -2.2(-10.3, 5.9) | 0.77 |
| PTS Di, % | -0.32(-5.0, 4.4) | 0.79 | -2.2(-10.8, 6.3) | 0.47 | 0.54(-7.3, 8.3) | 0.73 |
| RS Di, % | 0.25(-4.2, 4.7) | 0.70 | -0.58(-5.4, 4.3) | 0.73 | -0.74(-8.3, 6.8) | 0.69 |

Data are mean difference (95% limit of agreement (LOA)).
CS, circumferential strain; Di, dispersion; EDV, end-diastolic volume; ESV, end-systolic volume; EF, ejection fraction; GCS, global circumferential strain; GLS, global longitudinal strain; ICC, intra-class correlation coefficient; LS, longitudinal strain; LV, left ventricular; PTS, principle tangential strain; RS, radial strain; SDI, systolic dyssynchrony index; SV, stroke volume.

Test-retest reproducibility of 3D-STE derived measures was overall fair to good, but lower than when re-reading images were used as a measure of reliability (**Table-6**). Nevertheless, volumetric measures showed excellent reproducibility. All strain and rotational measures showed fair to good or excellent test-retest reproducibility, except for longitudinal strain and basal rotational measures, which showed fair and poor test-retest reproducibility respectively. SDIs showed good to excellent test-retest reproducibility, which was overall better than dispersion indices.

#### 3.2.2 Vascular Measurements

PWV, CACS and cIMT showed overall fair to good, excellent, and excellent reproducibility respectively. Test-retest reproducibility of PWV was done in 21 individuals. Mean difference (95% LOA) and ICC were −0.27 (−3.9, 3.3) m/s and 0.71 for PWV and 0.83 (−88.2, 89.9) mm and 0.54 for the carotid- femoral length measure. Repeatability and reproducibility of CACS were done in 20 individuals and were excellent. The within observer difference was –84.5 (−396.6, 565.7) and 0.99 ICC; and the between observer difference was –80.8 (−555.9, 394.3) and 0.99 ICC. Repeatability and reproducibility of cIMT and plaque characteristics (area and gray-scale median values) were excellent, while percentage white had fair to good repeatability and reproducibility (**Table-7**).

**Table.7.** Reproducibility of carotid intima medial thickness (cIMT) measures.

|  | Mean Diff (95% LOA) | ICC | Mean Diff (95% LOA) | ICC |
| --- | --- | --- | --- | --- |
|  | Intra-observer |  | Inter-observer |  |
| cIMT* far wall maximum of means, mm | 0.04(-0.08, 0.17) | 0.79 | -0.001(-0.12, 0.12) | 0.89 |
| Carotid lumen diameter, mm | 0.06(-0.16, 0.28) | 0.98 | 0.02(-0.46, 0.49) | 0.94 |
| Plaque area*, mm<sup>2</sup> | -3.3(-20.7, 14.1) | 0.81 | 0.3(-11.5, 12.1) | 0.90 |
| Gray-scale median, mm | 8.8(-13.4, 31.0) | 0.89 | 2.5(-57.2, 47.8) | 0.73 |
| Percentage white, % | 9.3(-13.4, 31.9) | 0.60 | 10.9(-16.6, 38.5) | 0.48 |

Data are mean difference (95% limit of agreement (LOA)). \*cIMT, $n=10$ ; plaque characteristics, $n=17$ . ICC, intra-class correlation coefficient.

## 4 Discussion

Over 1400 participants attended the SABRE 20 year follow-up and underwent a comprehensive health examination. The feasibility of image acquisition by most conventional echocardiographic measures was high. By contrast, 3DE had ∼76% acquisition feasibility. 3D-STE feasibility was highly influenced by image quality and only half of the datasets could be analyzed; these proportions are slightly better than reported by the Atherosclerosis Risk in Communities (ARIC) study (3, 22). The potential bias that may result from this missingness is an important consideration for future studies in older populations using 3D-STE (23–25).

Most conventional echocardiographic measurements and 3D-STE derived LV indices demonstrated good to excellent repeatability/reproducibility, in keeping with other studies (3, 26–31), although previous studies reporting the reproducibility of strain-based dyssynchrony measures by 3D-STE are scarce (14). Test-retest reproducibility, however, tended to be poorer for some measures, a factor that should be considered when performing longitudinal studies (26, 32).

We also show that vascular measures including cIMT, CACS and PWV are feasible in large cohort studies. The analyses of these measures are straightforward and reproducible and have been quite widely used. PWV, however, showed fair to good reliability, in keeping with previous studies (33).

SABRE is studying sizable proportions of first generation migrant African Caribbeans and South Asians, known to have different incident of CVD compared to White British (34). Capturing advanced and sensitive cardiovascular measures will enable new research opportunities related to CVD outcomes in this population, which we are currently investigating (35–38).

### 4.1 Limitations and Strengths

SABRE is the largest tri-ethnic UK cohort and includes a lengthy follow-up period with detailed phenotyping using state-of-the-art methods. A large percentage of participants could be traced 20 years later (∼90%) and 42% returned for clinic follow-up. However, in common with most longitudinal studies, elderly and/or infirm participants were more likely to decline to attend which may bias our feasibility estimates (**Table-S2**). 3DE was more frequently not feasible in older participants or those who were obese and diabetic, which may also introduce bias (39). Improvements in 3D probe technology may improve image quality future, although some scanning limitations related to morbidities such as atrial fibrillation, body habitus, or inability to lie on an echo couch are likely to remain problematic.

## 5 Conclusions

Capturing detailed cardiovascular phenotyping is reproducible and feasible in a large multi-ethnic study of older people. SABRE represents a uniquely rich resource of detailed information on cardiac and vascular structure and function in an ethnically diverse population and should lead to a better understanding of why people of South Asian and African Caribbean ancestry are at elevated risk of cardiometabolic diseases. The information reported in this study will be helpful to other research groups planning similar studies and can inform sample size calculations.

## Data Availability

The data underlying this article are available in the article and in its online supplementary material.

## Acknowledgments

LA is supported by a scholarship grant from Imam Abdulrahman Bin Faisal University for her postgraduate studies (PhD) at UCL. ADH and NC lead a unit that receives support from the UK MRC (Programme Code MC_UU_12019/1), they also receive support from BHF (PG/15/75/31748, CS/15/6/31468, CS/13/1/30327), and the National Institute for Health Research UCL Hospitals Biomedical Research Centre.

## References

1. Wild S, McKeigue P. Cross sectional analysis of mortality by country of birth in England and Wales, 1970–92. BMJ (Clinical research ed). 1997;314(7082):705–10.

2. Tillin T, Forouhi NG, McKeigue PM, Chaturvedi N, Group SS. Southall And Brent REvisited: Cohort profile of SABRE, a UK population-based comparison of cardiovascular disease and diabetes in people of European, Indian Asian and African Caribbean origins. Int J Epidemiol. 2012;41(1):33–42.

3. Shah AM, Cheng S, Skali H, Wu J, Mangion JR, Kitzman D, et al. Rationale and design of a multicenter echocardiographic study to assess the relationship between cardiac structure and function and heart failure risk in a biracial cohort of community-dwelling elderly persons: the Atherosclerosis Risk in Communities study. Circ Cardiovasc Imaging. 2014;7(1):173–81.

4. Mitchell C, Rahko PS, Blauwet LA, Canaday B, Finstuen JA, Foster MC, et al. Guidelines for Performing a Comprehensive Transthoracic Echocardiographic Examination in Adults: Recommendations from the American Society of Echocardiography. J Am Soc Echocardiogr. 2019;32(1):1–64.

5. Lang RM, Badano LP, Tsang W, Adams DH, Agricola E, Buck T, et al. EAE/ASE recommendations for image acquisition and display using three-dimensional echocardiography. Eur Heart J Cardiovasc Imaging. 2012;13(1):1–46.

6. Lang RM, Badano LP, Mor-Avi V, Afilalo J, Armstrong A, Ernande L, et al. Recommendations for Cardiac Chamber Quantification by Echocardiography in Adults: An Update from the American Society of Echocardiography and the European Association of Cardiovascular Imaging. J Am Soc Echocardiogr.. 2015;28(1):1–39.e14.

7. Du Bois D, Du Bois EF. A formula to estimate the approximate surface area if height and weight be known. 1916. Nutrition. 1989;5(5):303–11; discussion 12–3.

8. Dong SJ. Midwall fractional shortening in physiologic and pathologic left ventricular hypertrophy. Am J Hypertens. 2003;16(9 Pt 1):792–3.

9. de Simone G, Devereux RB, Roman MJ, Ganau A, Saba PS, Alderman MH, et al. Assessment of left ventricular function by the midwall fractional shortening/end-systolic stress relation in human hypertension. J Am Coll Cardiol. 1994;23(6):1444–51.

10. Nagueh SF, Smiseth OA, Appleton CP, Byrd BF, Dokainish H, Edvardsen T, et al. Recommendations for the Evaluation of Left Ventricular Diastolic Function by Echocardiography: An Update from the American Society of Echocardiography and the European Association of Cardiovascular Imaging. J Am Soc Echocardiogr.. 2016;29(4):277–314.

11. Brumback LC, Kronmal R, Heckbert SR, Ni H, Hundley WG, Lima JA, et al. Body size adjustments for left ventricular mass by cardiovascular magnetic resonance and their impact on left ventricular hypertrophy classification. Int J Cardiovasc Imaging. 2010;26(4):459–68.

12. Papachristidis A, Galli E, Geleijnse ML, Heyde B, Alessandrini M, Barbosa D, et al. Standardized Delineation of Endocardial Boundaries in Three-Dimensional Left Ventricular Echocardiograms. J Am Soc Echocardiogr. 2017;30(11):1059–69.

13. Tsang W, Kenny C, Adhya S, Kapetanakis S, Weinert L, Lang RM, et al. Interinstitutional measurements of left ventricular volumes, speckle-tracking strain, and dyssynchrony using three-dimensional echocardiography. J Am Soc Echocardiogr. 2013;26(11):1253–7.

14. Al Saikhan L, Park C, Hughes AD. Reproducibility of Left Ventricular Dyssynchrony Indices by Three-Dimensional Speckle-Tracking Echocardiography: The Impact of Sub-optimal Image Quality. Front Cardiovasc Med. 2019;6(149):149.

15. Laurent S, Cockcroft J, Van Bortel L, Boutouyrie P, Giannattasio C, Hayoz D, et al. Expert consensus document on arterial stiffness: methodological issues and clinical applications. Eur Heart J. 2006;27(21):2588–605.

16. Vlachopoulos C, Aznaouridis K, O’Rourke MF, Safar ME, Baou K, Stefanadis C. Prediction of cardiovascular events and all-cause mortality with central haemodynamics: a systematic review and meta-analysis. Eur Heart J. 2010;31(15):1865–71.

17. Chen CH, Nevo E, Fetics B, Pak PH, Yin FC, Maughan WL, et al. Estimation of central aortic pressure waveform by mathematical transformation of radial tonometry pressure. Validation of generalized transfer function. Circulation. 1997;95(7):1827–36.

18. Wendelhag I, Liang Q, Gustavsson T, Wikstrand J. A new automated computerized analyzing system simplifies readings and reduces the variability in ultrasound measurement of intima-media thickness. Stroke. 1997;28(11):2195–200.

19. Stein JH, Korcarz CE, Hurst RT, Lonn E, Kendall CB, Mohler ER, et al. Use of carotid ultrasound to identify subclinical vascular disease and evaluate cardiovascular disease risk: a consensus statement from the American Society of Echocardiography Carotid Intima-Media Thickness Task Force. Endorsed by the Society for Vascular Medicine. J Am Soc Echocardiogr. 2008;21(2):93–111; quiz 89–90.

20. Hughes AD, Falaschetti E, Witt N, Wijetunge S, Thom SA, Tillin T, et al. Association of Retinopathy and Retinal Microvascular Abnormalities With Stroke and Cerebrovascular Disease. Stroke. 2016;47(11):2862–4.

21. Rosner B. Fundamentals of Biostatistics. Cengage Learning, Inc; 7th edition, 2010.

22. Hung CL, Goncalves A, Shah AM, Cheng S, Kitzman D, Solomon SD. Age-and Sex-Related Influences on Left Ventricular Mechanics in Elderly Individuals Free of Prevalent Heart Failure: The ARIC Study (Atherosclerosis Risk in Communities). Circ Cardiovasc Imaging. 2017;10(1).

23. Al Saikhan L, Park C, Hardy R, Hughes A. Prognostic implications of left ventricular strain by speckle-tracking echocardiography in population-based studies: a systematic review protocol of the published literature. BMJ Open. 2018;8.

24. Al Saikhan L, Park C, Hardy R, Hughes A. Prognostic implications of left ventricular strain by speckle-tracking echocardiography in the general population: a meta-analysis. Vasc Health Risk Manag. 2019;15:229–51.

25. Al Saikhan L, Park C, Hughes A. P644 The impact of intentional distortion of image quality on left ventricular deformation indices by three-dimensional speckle-tracking echocardiography. Eur Heart J Cardiovasc Imaging. (2019) 20(Suppl. 1):i363–i81.

26. Kaku K, Takeuchi M, Tsang W, Takigiku K, Yasukochi S, Patel AR, et al. Age-Related Normal Range of Left Ventricular Strain and Torsion Using Three-Dimensional Speckle-Tracking Echocardiography. J Am Soc Echocardiog. 2014;27(1):55–64.

27. Muraru D, Cucchini U, Mihăilă S, Miglioranza MH, Aruta P, Cavalli G, et al. Left Ventricular Myocardial Strain by Three-Dimensional Speckle-Tracking Echocardiography in Healthy Subjects: Reference Values and Analysis of Their Physiologic and Technical Determinants. J Am Soc Echocardiog. 2014;27(8):858–71.e1.

28. Martinez C, Ilardi F, Dulgheru R, Sugimoto T, Bernard A, Lancellotti P, et al. 3D echocardiographic reference ranges for normal left ventricular volumes and strain: results from the EACVI NORRE study. Eur Heart J Cardiovasc Imaging. 2016;18(4):475–83.

29. Kleijn SA, Aly MF, Knol DL, Terwee CB, Jansma EP, Abd El-Hady YA, et al. A meta-analysis of left ventricular dyssynchrony assessment and prediction of response to cardiac resynchronization therapy by three-dimensional echocardiography. Eur Heart J Cardiovasc Imaging. 2012;13(9):763–75.

30. Russo C, Jaubert MP, Jin Z, Homma S, Di Tullio MR. Intra-and interobserver reproducibility of left ventricular mechanical dyssynchrony assessment by real time three-dimensional echocardiography. Echocardiography. 2012;29(5):598–607.

31. Yuda S, Sato Y, Abe K, Kawamukai M, Kouzu H, Muranaka A, et al. Inter-vendor variability of left ventricular volumes and strains determined by three-dimensional speckle tracking echocardiography. Echocardiography. 2014;31(5):597–604.

32. Al Saikhan L, Park C, Hughes A. Image Quality Causes Substantial Bias in Three-Dimensional Speckle-Tracking Echocardiography Measures. bioRxiv. 2018:500777. doi.org/10.1101/500777

33. Meyer ML, Tanaka H, Palta P, Patel MD, Camplain R, Couper D, et al. Repeatability of Central and Peripheral Pulse Wave Velocity Measures: The Atherosclerosis Risk in Communities (ARIC) Study. Am J Hypertens. 2015;29(4):470–5.

34. Tillin T, Hughes AD, Mayet J, Whincup P, Sattar N, Forouhi NG, et al. The relationship between metabolic risk factors and incident cardiovascular disease in Europeans, South Asians, and African Caribbeans: SABRE (Southall and Brent Revisited) –-a prospective population-based study. J Am Coll Cardiol. 2013;61(17):1777–86.

35. Al Saikhan L, Park C, Tillin T, Mayet J, Chaturvedi N, Hughes A. 3 3D echocardiography-derived indices of left ventricular function and structure predict long-term mortality differently in men and women: the Southall And Brent Revisited (SABRE) study. Heart. 2019;105:A3–A5.

36. Al Saikhan L, Park C, Tillin T, Williams S, Mayet J, Chaturvedi N, et al. P2444Comparison of 3D and 2D echocardiography-derived indices of left ventricular function and structure to predict long-term mortality in the general population: Southall And Brent Revisited (SABRE) study. Eur Heart J. 2019;40(Supplement_1).

37. Al Saikhan L, Park C, Tillin T, Mayet J, Chaturvedi N, Hughes A. 3.5 Arterial Stiffness Partly Explains Sex Differences in Associations Between Left Ventricular Structure and Mortality: The Southall and Brent Revisited (Sabre) Study. Artery Research. 2020.

38. Al Saikhan L, Park C, Tillin T, Mayet J, Chaturvedi N, Hughes A. THE PREVALENCE OF SUBCLINICAL LEFT VENTRICULAR SYSTOLIC DYSFUNCTION BY 3D-GLOBAL LONGITUDINAL STRAIN IN SOUTH ASIANS COMPARED TO WHITES EUROPEANS AND THE ROLE OF CENTRAL OBESITY: THE SOUTHALL AND BRENT REVISITED (SABRE) STUDY. J Am Coll Cardiol. 2020;75(11 Supplement 1):1656.

39. Park CM, March K, Ghosh AK, Jones S, Coady E, Tuson C, et al. Left-ventricular structure in the Southall And Brent REvisited (SABRE) study: explaining ethnic differences. Hypertension. 2013;61(5):1014–20.

